# High-sensitivity C-reactive protein modifies the prognostic value of platelet count for clinical outcomes after ischemic stroke

**DOI:** 10.1101/2023.02.27.23286541

**Authors:** Fanghua Liu, Pinni Yang, Yinan Wang, Mengyao Shi, Ruirui Wang, Qingyun Xu, Yanbo Peng, Jing Chen, Jintao Zhang, Aili Wang, Tan Xu, Yong-hong Zhang, Jiang He

## Abstract

**Background:** Platelets play a critical role in the formation of thrombosis and embolism, and high-sensitivity C-reactive protein (HS-CRP) is an important indicator of inflammation, which contribute to the development of ischemic stroke development. This study aimed to examine whether the relationship between baseline platelet count and adverse clinical outcomes is modulated by HS-CRP in patients with ischemic stroke.

**Methods:** A total of 3267 patients with ischemic stroke were included in the analysis. The primary outcome was a combination of death and major disability (modified Rankin Scale [mRS] score ≥3) at 1 year after ischemic stroke. Secondary outcomes included major disability, death, vascular events, composite outcome of vascular events or death, and an ordered 7-level categorical score of the mRS at 1 year. Multivariate logistic regression and Cox proportional hazards regression models were used to assess the association between the baseline platelet count and clinical outcomes stratified by HS-CRP levels when appropriate, odds ratio (OR) or hazard ratio (HR) and 95% confidence interval (CI) were calculated for the highest quartiles of platelet counts compared with the lowest quartile.

**Results:** The elevated platelet count was associated with the primary outcome (OR, 3.14;95% CI, 1.77-5.58), major disability (OR, 2.07;95%CI, 1.15-3.71), death (HR, 2.75;95%CI, 1.31-5.79), composite outcome of vascular events or death (HR, 2.57;95%CI, 1.38-4.87), and the ordered 7-level categorical score of the mRS at 1 year (OR, 2.01 [95%CI, 1.33-3.04]) among patients with high HS-CRP levels (all *P* _trend_ <0.05). However, platelet count was not associated with the primary outcome (OR, 1.13; 95%CI, 0.75-1.71), major disability (OR, 1.34; 95%CI, 0.87-2.08), death (HR, 0.48; 95%CI, 0.19-1.24), composite outcome of vascular events or death (HR, 0.79; 95%CI, 0.45-1.37), and the ordered 7-level categorical score of the mRS at 1 year (OR, 1.17; 95%CI, 0.91-1.49) (all *P* _trend_ >0.05) in those with low HS-CRP levels. There was an interaction effect of platelet count and HS-CRP on the primary outcome, death, composite outcome of vascular events or death, and the ordered 7-level categorical score of the mRS at 1 year after ischemic stroke (all *P* _interaction_ <0.05).

**Conclusions:** An elevated platelet count was associated with adverse clinical outcomes in ischemic stroke patients with high HS-CRP levels but not in those with low HS-CRP levels, the HS-CRP level had a modifying effect on the association between platelet count and clinical outcomes in patients with ischemic stroke, suggesting that strategies for anti-inflammatory and antiplatelet therapy should be developed according to the results of both platelet and HS-CRP testing.

## Introduction

Stroke is still the second leading cause of mortality and the main cause of disability worldwide, with 12.2 million incident stroke cases in 2019^1^, leading to a serious disease burden in low-and middle-income countries^2^. Ischemic stroke accounts for >70% of incident strokes^3, 4^. Early and economic risk assessment, including estimates of disease severity and identification of modifiable risk factors for unfavorable outcomes, may be helpful for the secondary prevention of stroke^5^.

Thrombosis and inflammatory reactions are important mechanisms in the occurrence and development of ischemic stroke^6-10^. Platelets play a critical role in the formation of thrombosis and embolism, and high-sensitivity C-reactive protein (HS-CRP) is an important indicator of inflammation, which contributes to the development of cardio-cerebrovascular atherosclerosis and the initiation of ischemic stroke development^11-17^. It has been reported that increased platelet counts are associated with cardiovascular events and mortality in the general population^18, 19^. In a Mendelian randomization study, a higher genetically determined platelet count increased the risk of ischemic stroke^20^. However, the relationship between platelet count and ischemic stroke prognosis remains inconclusive, and two studies^21, 22^ showed a significant association of increased platelet count with the development of functional outcomes in ischemic stroke, while one study showed no significant association^23^. Mounting evidence suggests that an elevated HS-CRP level reflects the instability of atherosclerotic plaques^14^ and is associated with the risk of cardiovascular diseases, stroke, and adverse clinical outcomes after stroke^14-17, 24^. Elevated HS-CRP levels have been reported to increase plasma von Willebrand factor levels^25^, which in turn affect the activity of platelets^12, 26^. However, the effect of HS-CRP on the association between platelet count and clinical outcomes of ischemic stroke is unclear. Thus, the aim of the present study was to evaluate the prognostic value of platelet count in patients with ischemic stroke stratified by HS-CRP levels.

## Methods

### Study participants

This study was conducted based on the China Antihypertensive Trial in Acute Ischemic Stroke (CATIS), a multicenter, single-blind, blinded endpoint randomized clinical trial that was conducted in 26 hospitals across China. More details on the rational design and major results of the CATIS trial have been reported in previous publications^27^. Briefly, 4071 patients were recruited for the CATIS. The inclusion criteria for the CATIS were as follows: (1) age ≥22 years, (2) ischemic stroke confirmed by computed tomography or magnetic resonance imaging of the brain within 48 h of symptom onset, and (3) elevated systolic blood pressure (BP) between 140 mmHg and <220 mmHg. The exclusion criteria were as follows: (1) systolic BP ≥220 or diastolic BP ≥120 mmHg, (2) severe heart failure, (3) acute myocardial infarction or unstable angina, (4) atrial fibrillation, (5) aortic dissection, (6) cerebrovascular stenosis (≥70%), (7) resistant hypertension, (8) deep coma, and (9) treatment with intravenous thrombolytic therapy. Furthermore, 804 patients were excluded due to a lack of baseline HS-CRP or platelet count recordings (N=644) or loss to follow-up at 1 year (N=160). Finally, 3267 patients with acute ischemic stroke were included in the present analysis (**Figure 1**).

**Figure 1.**
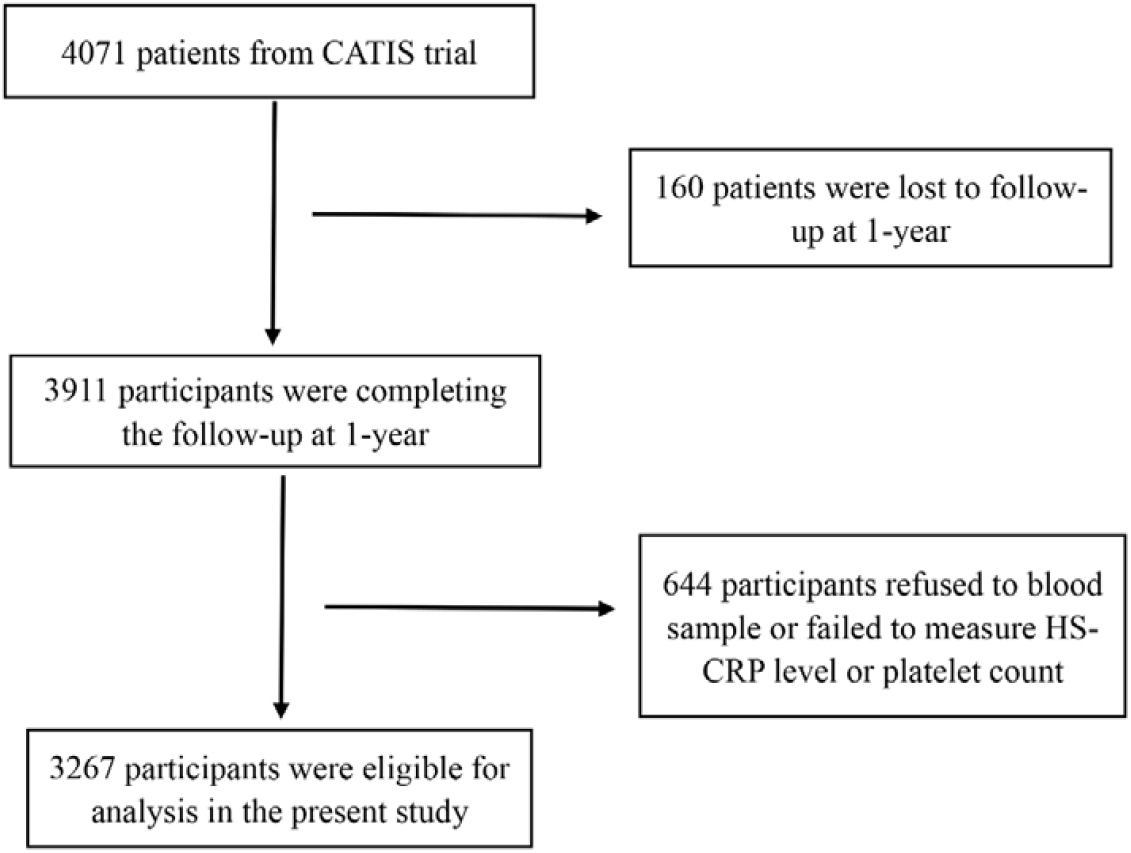
Study participants flow char.

This study was approved by the Institutional Review Boards of Soochow University in China and Tulane University in the United States, as well as by the ethical committees of the 26 participating hospitals. Written informed consent was obtained from all study participants or their immediate family members.

### Data collection and measurements

Baseline data on demographic characteristics, medication history, and clinical features were collected at enrollment. The National Institutes of Health Stroke Scale (NIHSS) was used to evaluate the stroke severity^28^. According to the symptoms and imaging data of the patients, ischemic stroke was classified as large-artery atherosclerosis, cardiac embolism, and small-artery occlusion lacunae (lacunar)^29^. Three baseline BP measurements were obtained by trained nurses while the patient was in the supine position using a standard mercury sphygmomanometer^30^. The mean of the three BP records was used in the analyses.

Blood samples were collected within 24 hours of hospital admission after at least 8 hours of fasting. Routine laboratory analyses (those of blood glucose, blood lipids, platelet count, etc.) were performed for all enrolled patients at each participating hospital upon admission. Body weight and height were measured using standard methods, and the body mass index was calculated as weight in kilograms divided by the square of height in meters (kg/m^2^). The HS-CRP concentration was measured using commercially available immunoassays (R&D Systems). Dyslipidemia was defined as total cholesterol more than 5.2 mmol/L, triglycerides more than 1.7 mmol/L, low-density lipoprotein cholesterol more than 3.4 mmol/L, or high-density lipoprotein cholesterol less than 1.0 mmol/L according to Chinese guidelines on the prevention and treatment of dyslipidemia in adults^31^. The estimated glomerular filtration rate was calculated using the Chronic Kidney Disease Epidemiology Collaboration creatinine equation with an adjusted coefficient of 1.1 for the Chinese population^32^.

### Outcome assessment

Participants were followed up at 1 year after stroke by trained neurologists unaware of random assignment^27^. The primary outcome was a 1-year poor functional outcome (mRS score, 3-6). The secondary outcomes were major disability (mRS score of 3-5), death (mRS score of 6), vascular events (i.e., recurrent nonfatal stroke, nonfatal myocardial infarction, hospitalized and treated angina, hospitalized and treated congestive heart failure, and hospitalized and treated peripheral arterial disease), and a composite outcome of vascular events or death. We further included an ordered 7-level categorical score of the mRS at 1 year as an outcome of the neurologic functional status^33^. The death certificates were obtained for all deceased participants, and hospital data were abstracted for vascular events. The study outcome assessment committee, blinded to random assignment, reviewed and adjudicated subsequent outcomes based on the criteria established in the Antihypertensive and Lipid-Lowering Treatment to Prevent Heart Attack Trial (ALLHAT)^34^.

### Statistical analysis

First, the patients were divided into low HS-CRP level (<4.8 mg/L) and high HS-CRP level (≥4.8 mg/L) subgroups based on the cut-off value of the third quartile of baseline HS-CRP levels. Second, the platelet count was divided into four groups according to the baseline platelet count quartiles within each HS-CRP subgroup. Tests for linear trends in baseline characteristics across the platelet count quartiles in each HS-CRP subgroup were performed using generalized linear regression analysis for continuous variables and the Cochran-Armitage trend Chi-square test for categorical variables. The baseline characteristics of the patients included in the present study and all patients enrolled in the CATIS were compared using Student’s t-test, Wilcoxon rank-sum test, or the Chi-square test as appropriate (**Supplemental Table S1**).

Multivariate logistic regression and Cox proportional hazards regression models were used to assess the association between the baseline platelet count and clinical outcomes stratified by HS-CRP levels when appropriate. Odds ratios (ORs) or hazard ratios (HRs) and 95% confidence intervals (CI) were calculated for the upper quartiles of platelet counts compared with the lowest quartile. Model 1 was adjusted for age and gender. Model 2 was further adjusted for current cigarette smoking, current alcohol drinking, the NIHSS score at admission, time to randomization after admission, body mass index, randomized antihypertensive status, dyslipidemia, estimated glomerular filtration rate, history of hypertension, family history of stroke, history of diabetes mellitus, use of lipid-lowering drugs, coronary heart disease, blood glucose, white blood cell counts, systolic BP at baseline, and the ischemic stroke subtype based on Model 1. Model 3 was further adjusted for antiplatelet therapy after admission based on Model 2. The effect of the interaction between the HS-CRP level and platelet count on study outcomes was also tested using the likelihood ratio test in models with multiplicative interaction terms. Nonparametric restricted cubic splines were used to explore the shape of the association between the platelet count and adverse clinical outcomes after ischemic stroke with four knots (at the 5th, 35th, 65th, and 95th percentiles of the subgroup-specific distribution of platelet counts)^35^.

To test the robustness of our findings, thrombocytopenia (platelet count <100 × 10^9^/L, N=79) and thrombocytosis (platelet count >450 × 10^9^/L, N=10) were further excluded from the sensitivity analyses conducted in multivariate adjusted logistic regression models or multivariate adjusted Cox proportional hazards models.

All *P* values were two tailed, and a significance level of 0.05 was used. Statistical analyses were performed using SAS statistical software (version 9.4, Cary, NC, USA).

## Results

### Baseline characteristics

The baseline characteristics of the patients included in this study were balanced with those of the total CATIS population (**Supplemental Table S1**). A total of 3267 individuals (2093 men and 1174 women) with an average age of 62.4 years were included. Among the patients with high HS-CRP levels, those with higher platelet counts tended to be female, had a lower rate of drinking and smoking, had a higher prevalence of diabetes mellitus, and had higher low-density lipoprotein cholesterol, total cholesterol, and blood glucose levels and white blood cell counts. Among the patients with low HS-CRP levels, those with higher platelet counts were more likely to be younger and female and have lower SBP levels, lower rates of alcohol consumption and diabetes mellitus, higher white blood cell counts and triglyceride, total cholesterol, low-density lipoprotein cholesterol, and high-density lipoprotein cholesterol levels, and a longer time from onset to randomization (**Table 1**).

**Table 1.**
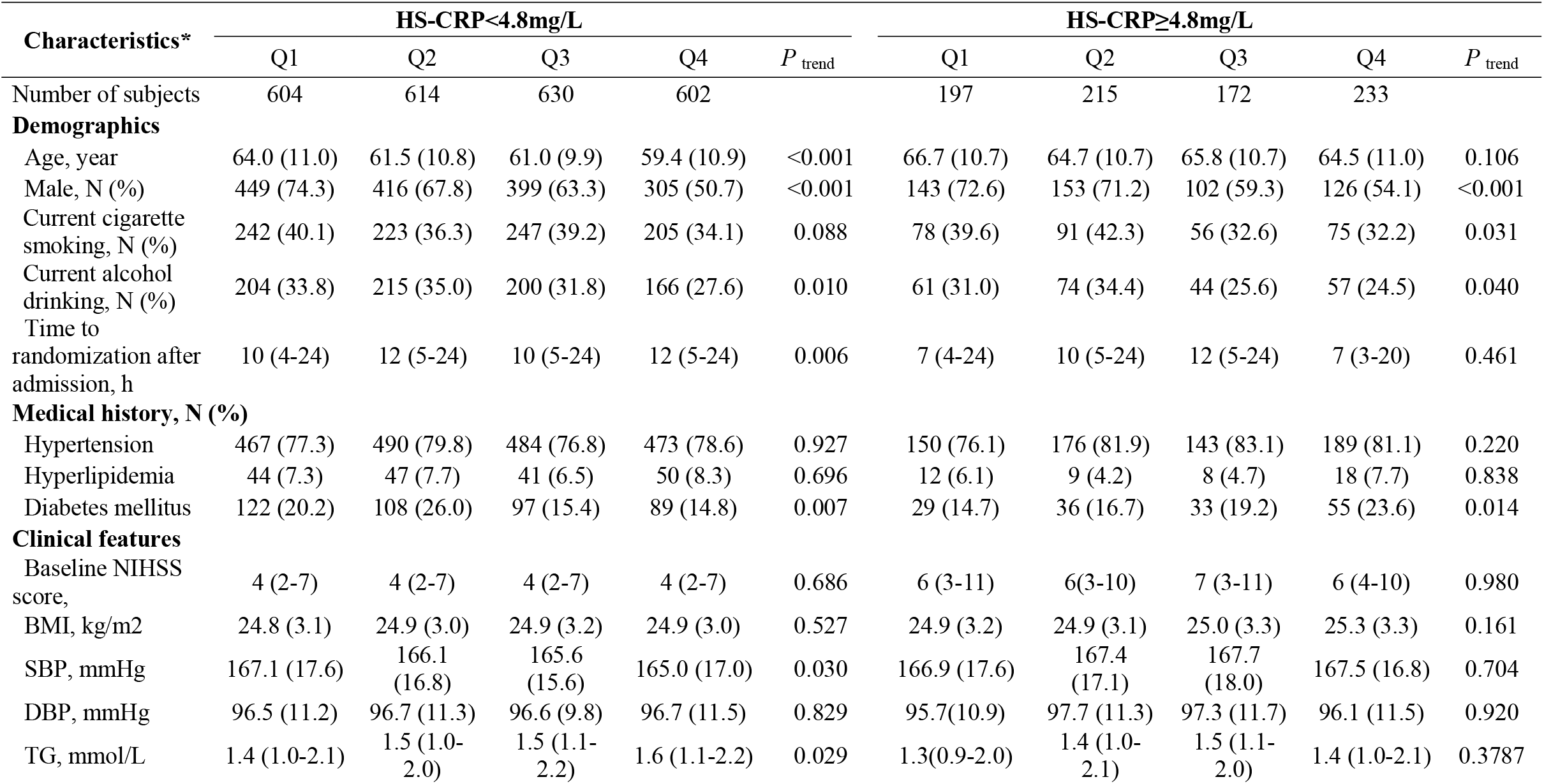
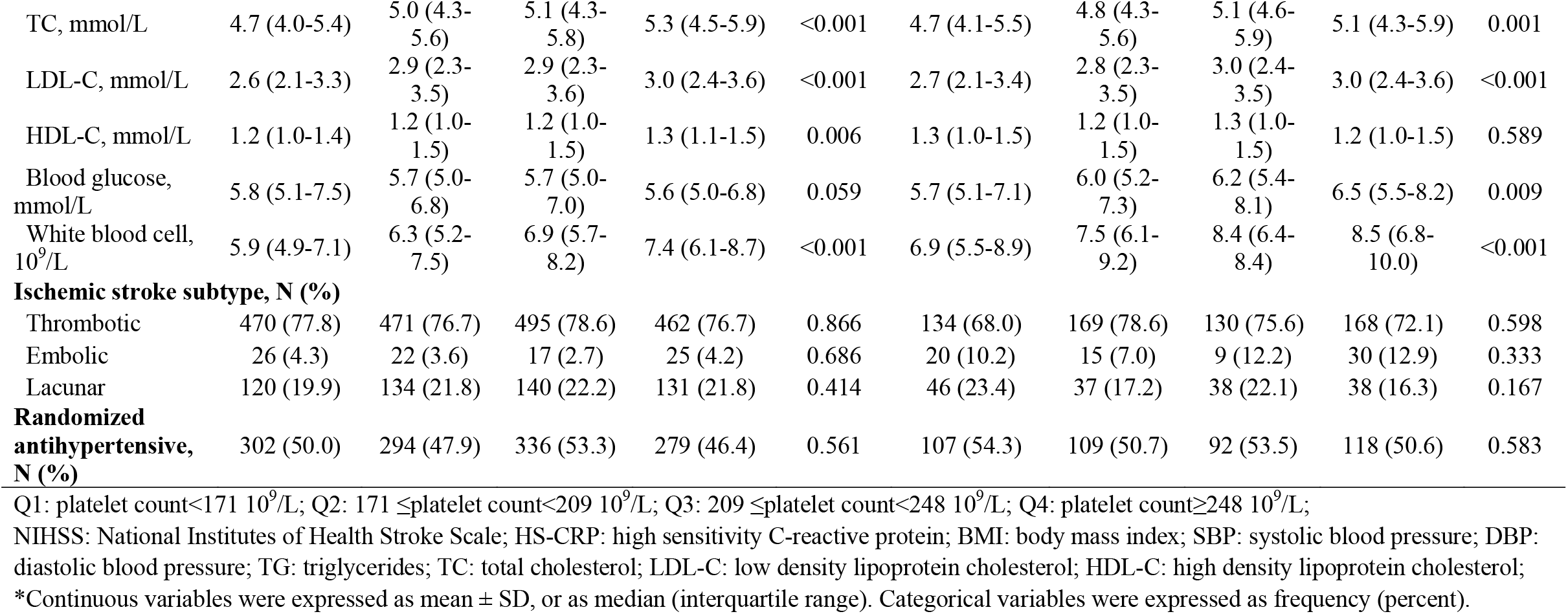
Baseline characteristics of study participants with different baseline platelet counts by stratified by HS-CRP levels.

### Baseline platelet count and clinical outcomes

Within 1 year after ischemic stroke, 419 (17.10%) patients in the low HS-CRP group and 317 (38.80 %) patients in the high HS-CRP group met the primary endpoint of poor functional outcomes. In the high HS-CRP group, the multivariable-adjusted ORs (95%CI) or HRs (95%CI) for the primary outcome, major disability, death, composite outcome of vascular events or death, and the ordered 7-level categorical score of the mRS at 1 year for patients in the highest quartile of platelet count compared with the lowest quartile of platelet count were 3.14 (1.77-5.58), 2.07 (1.15-3.71), 2.75 (1.31-5.79), 2.63 (1.32-5.21), and 1.97 (1.28-3.03), respectively, while the multivariable-adjusted ORs (95%CI) or HRs (95%CI) for the primary outcome, major disability, death, composite outcome of vascular events or death, and the ordered 7-level categorical score of the mRS at 1 year for patients in the highest quartile of platelet count compared with the lowest quartile were 1.13(0.75-1.71), 1.34 (0.87-2.08), 0.48 (0.19-1.24), 0.79(0.45-1.37), and 1.17 (0.91-1.49) (all *P* _trend_ >0.05) in those with low HS-CRP levels, which showed an interaction effect of the platelet count and HS-CRP on the primary outcome (*P* _interaction_ =0.002), death (*P* _interaction_ =0.002), composite outcome of vascular events or death (*P* _interaction_ =0.016), and the ordered 7-level categorical score of the mRS at 1 year (*P* _interaction_ =0.035), HS-CRP significantly modified the prognostic value of the platelet count for the clinical outcomes of ischemic stroke (**Table 2**). After further exclusion of thrombocytopenia and thrombocytosis, our findings were consistent with the original results, confirming the robustness of our results (**Table 3**).

**Table 2.**
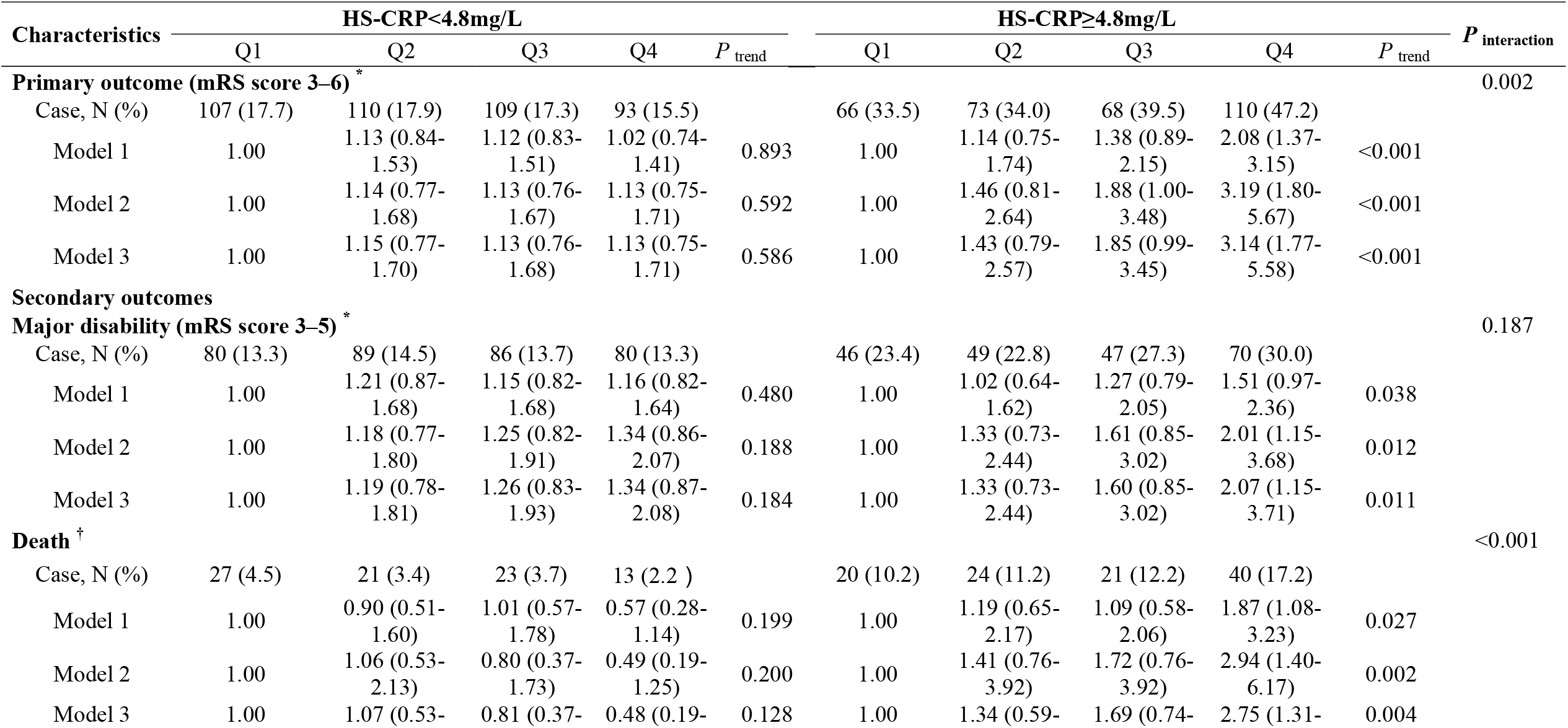
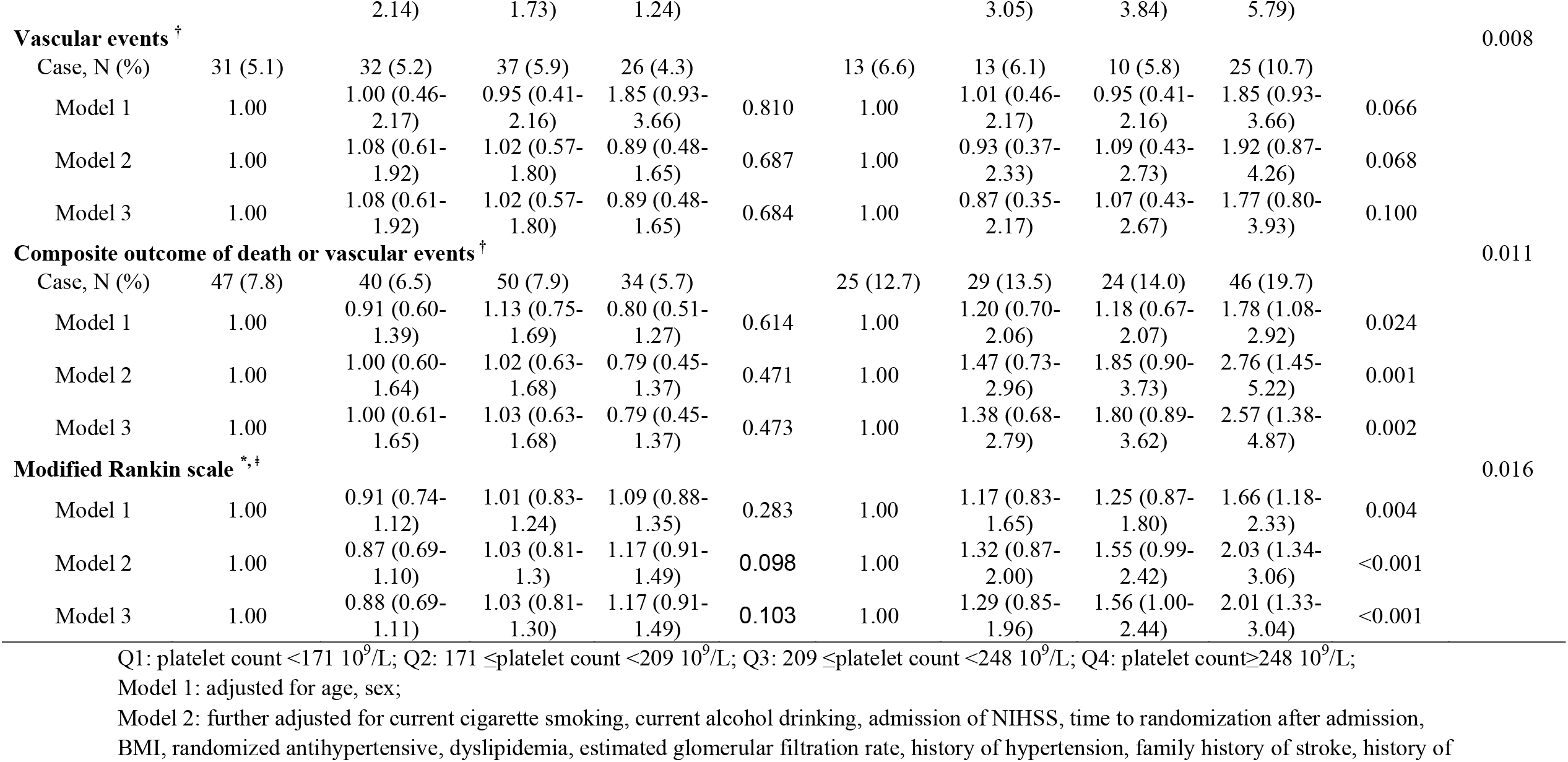
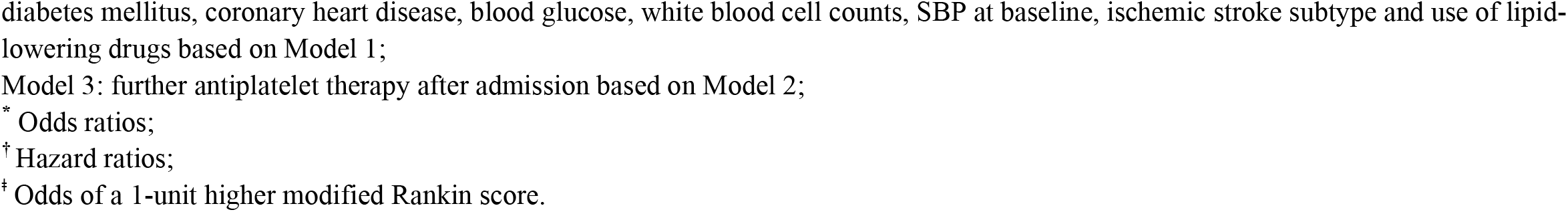
Odds ratios or Hazard ratios and 95% confidence intervals of outcome for quartile of baseline platelet counts by stratified by HS-CRP levels at baseline.

**Table 3.**
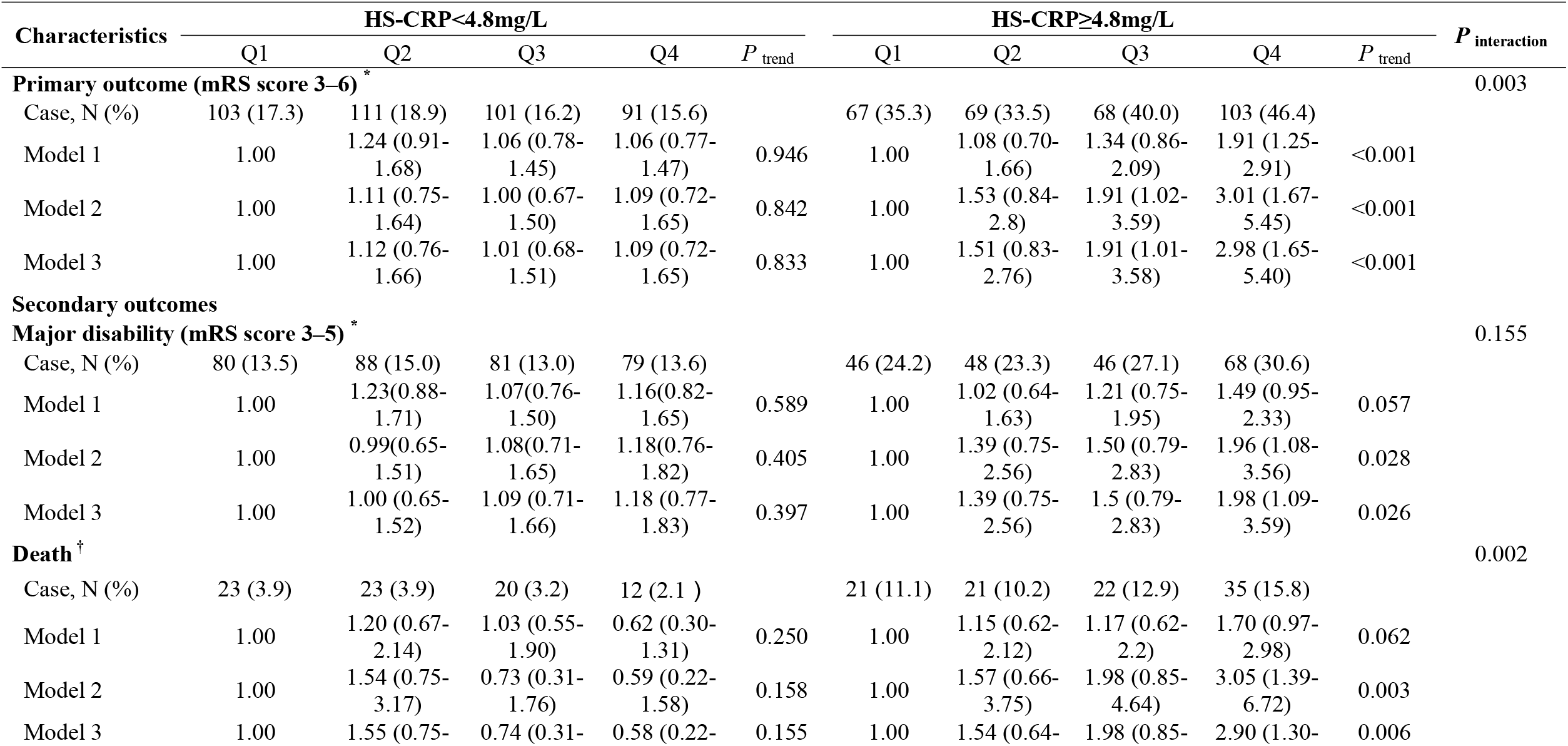
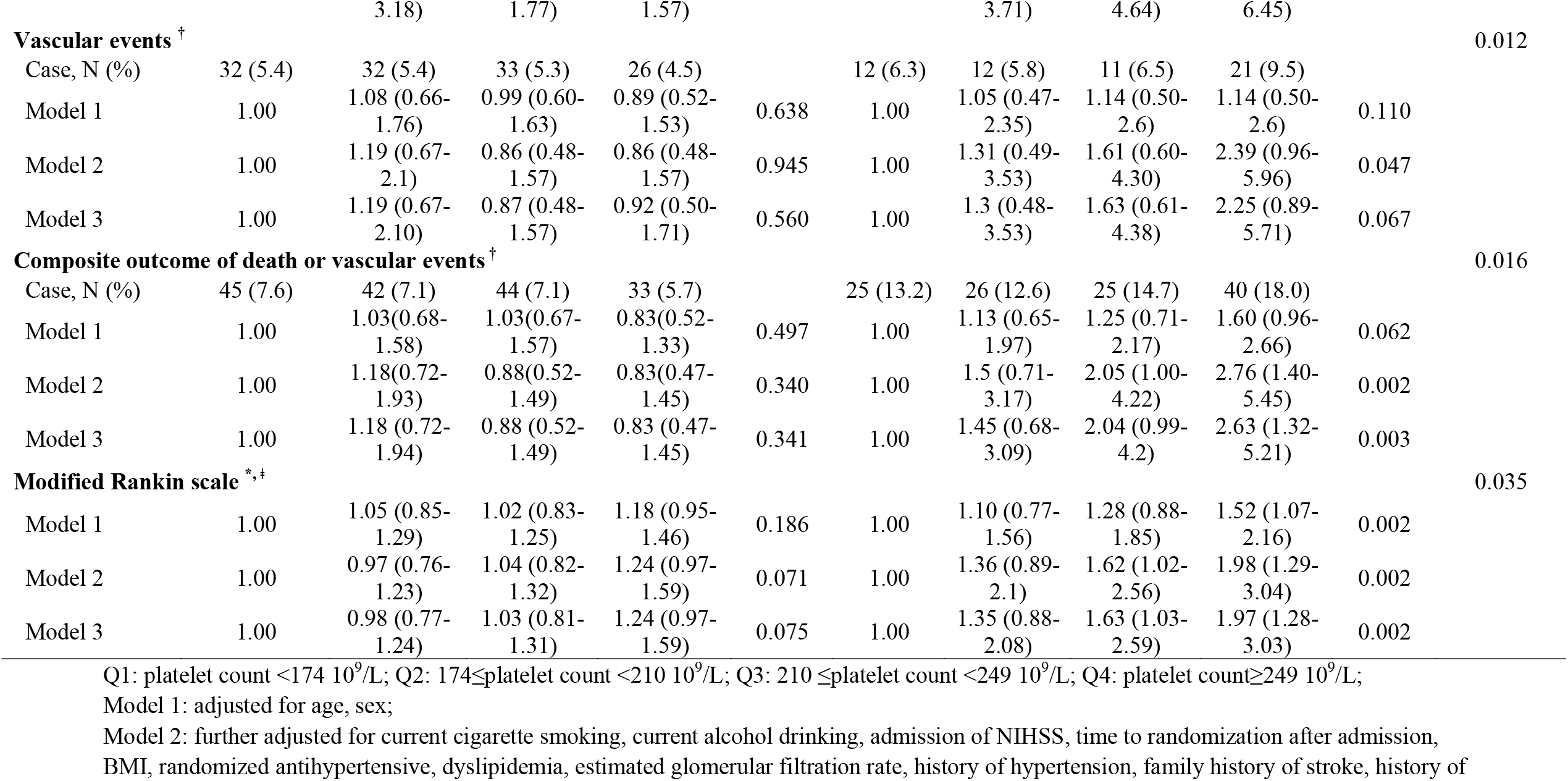
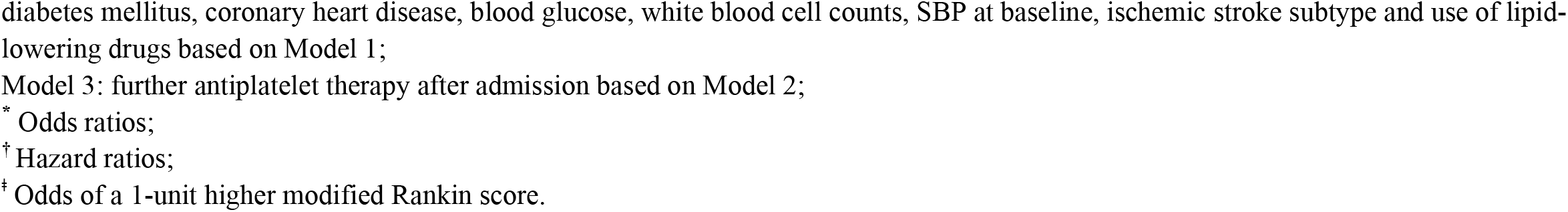
Odds ratios or Hazard ratios and 95% confidence intervals of study outcomes for quartiles of baseline platelet count stratified by HS-CRP level after excluding thrombocytopenia and thrombocytosis.

Multivariate adjusted spline regression models showed a linear association between the platelet count and primary outcome (*P* _linearity_ <0.001) and composite outcome of vascular events or death (*P* _linearity_ <0.001) in the high HS-CRP group (**Figure 2, [B], [D]**). Moreover, there was a dose-dependent relationship between the platelet count and 1-year mRS score (*P* _trend_ <0.001, **Figure 3 and Table 2**).

**Figure 2.**
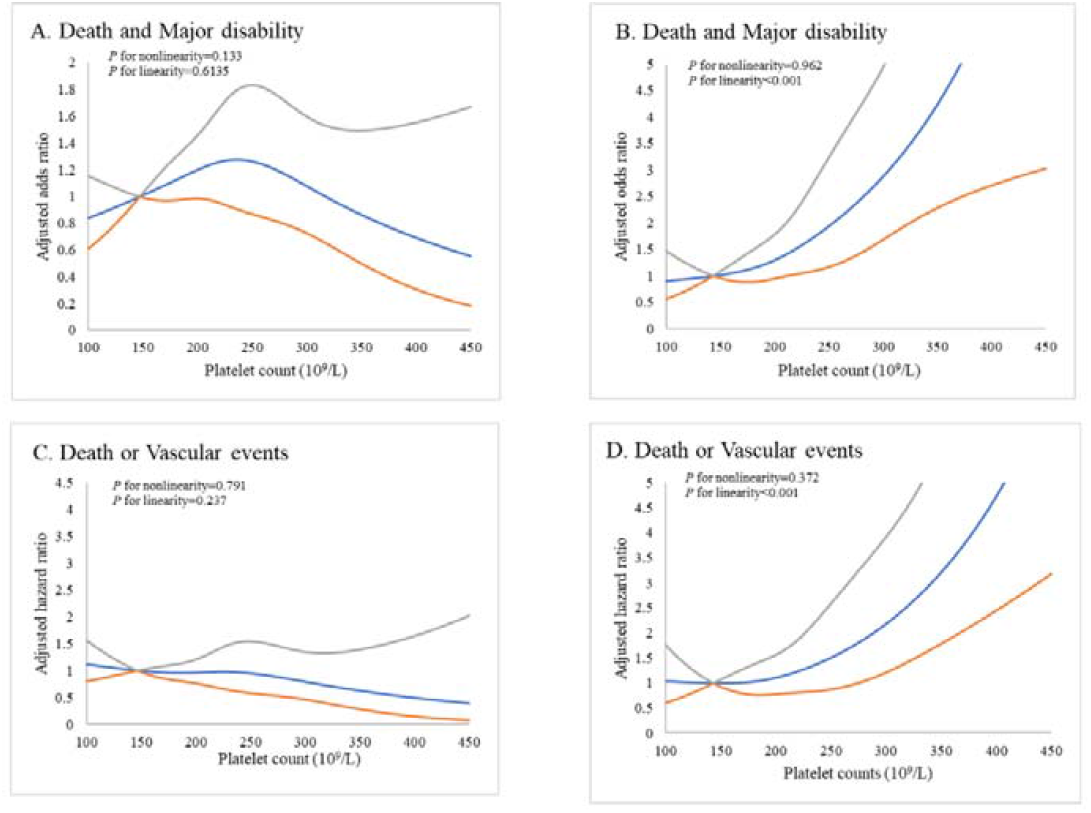
Linear test of the association between baseline blood platelet count and 1-year clinical outcomes. Odds ratio or hazard ratio and 95% confidence interval derived from restricted cubic spline regression, with knots placed at the 5th, 35th, 65th, and 95th percentiles of platelet count. Odds ratios or hazard ratios were adjusted for the same variables as model 3 in Table 2. (A) HS-CRP<4.8 mg/L; (B) HS-CRP≥4.8mg/L; (C) HS-CRP<4.8 mg/L; (D) HS-CRP≥4.8mg/L.

**Figure 3.**
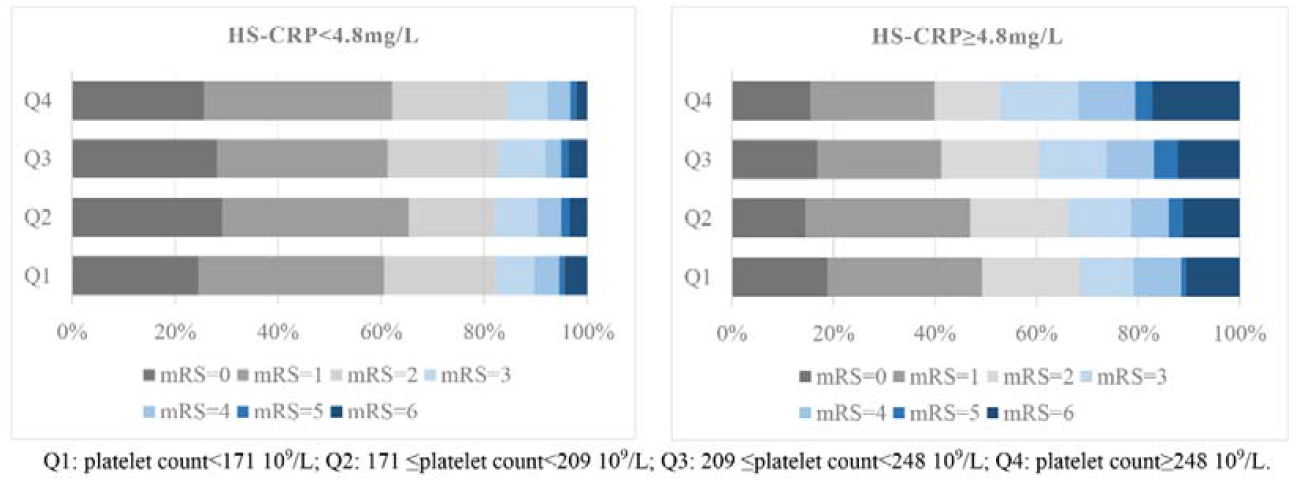
Baseline platelet count and 1-year mRS score. Multivariable-adjusted odds ratio of ordinal logistic regression analysis was 2.03(95% CI, 1.34-3.06) for patients in the highest quartile of platelet count compared with the patients in the lowest quartile in high HS-CRP group (*P* _trend_<0.001). Multivariable model adjusted for the same variables as model 3 in Table.

## Discussion

To our knowledge, this is the first large prospective study to assess the prognostic value of the baseline platelet count for clinical outcomes according to HS-CRP stratification among patients with ischemic stroke. In the present study based on the CATIS, we found that an elevated platelet count was independently associated with an increased risk of primary outcome, death, and composite outcome of vascular events or death, and the ordered 7-level categorical score of the mRS within one year after ischemic stroke among patients with high HS-CRP levels, but not in those with low HS-CRP levels. Thus, these results strongly support the theory that high HS-CRP levels could significantly modify the prognostic value of the platelet count for clinical outcomes after ischemic stroke and provide valuable insights into adding anti-inflammatory therapy to the secondary prevention of ischemic stroke^36^ while administering antiplatelet therapy.

Previous studies have reported the involvement of platelets in maintaining vascular homeostasis and mediating immune responses, inflammation, and atherosclerosis^11^. In an observational study, the platelet count showed a U-shaped relationship with future cardiovascular events and mortality in the general population^18^. However, the relationship between the platelet count and stroke prognosis has been inconsistent. In a cohort of 16,842 participants with stroke, platelet counts within normal ranges were associated with long-term stroke recurrence, mortality, and poor functional outcome^21^. In contrast, Du et al^37^ reported that an elevated platelet count was not significantly associated with prognosis 30 days after ischemic stroke. Recently, a study investigated the prognostic value of the platelet count for clinical outcomes in patients with ischemic stroke and transient ischemic attack. Patients in the top quintile of platelet count (249–450 × 10^9^/L) had an increased risk of death (adjusted HR=1.43, 95% CI 1.19–1.73) and poor functional outcome (adjusted OR = 1.49, 95% CI=1.28–1.74) at 1-year follow-up compared to those in the low platelet count quintile (186–212 × 10^9^/L)^21^. However, in a prospective study of 553 patients with first-ever ischemic stroke, Ghodsi et al. reported that increased mean platelet volume, not platelet count, was significantly associated with mortality within 3 months (OR=3.88, 95%CI=2.04-7.38) and 1 year (OR=3.32, 95%CI=1.91-5.78), as well as poor function at 3 months (OR=3.25, 95%CI=1.80-5.86) and 1 year (OR=4.35, 95%CI=2.36-8.02)^23^. Among the above studies, some included transient ischemic attack (TIA)^37^ or used small samples^23, 37^, which did not consider the effect of HS-CRP on the relationship between the platelet count and clinical outcome of stroke. Both thrombosis and inflammatory reactions are important mechanisms for the occurrence and development of ischemic stroke^14, 38, 39^, which may affect the prognosis after stroke in the form of their interaction. Our study found a linear relationship between the baseline platelet count and poor outcomes of ischemic stroke only in patients with high HS-CRP levels, but not in those with low HS-CRP levels, and the association persisted in a sensitivity analysis in which patients with thrombocytopenia or thrombocytosis were excluded. Our results suggest that HS-CRP modifies the prognostic value of the platelet count for the clinical outcome of ischemic stroke.

As an inflammatory factor, HS-CRP is considered an important marker of atherosclerotic rupture^14, 38, 39^ and is associated with incidence risk^17, 24^ and poor prognosis risk of ischemic stroke^40, 41^. Several mechanisms may underlie the observed interaction between the HS-CRP level and platelet count in the prognosis of ischemic stroke. The platelet count and HS-CRP level are important indicators of thrombosis and inflammation in the development of ischemic stroke^13, 17^. Moreover, several studies have found that elevated HS-CRP levels not only promote an increase in the plasma von Willebrand factor concentration^25^ but also stimulate the expression of MMP-2 and MMP-9^14, 38, 39^, which are directly involved in the rupture of atherosclerotic plaques. Ruptured plaques can promote the adhesion of von Willebrand factor and reduce platelet velocity, thereby stimulating the activation of platelet function, enhancing platelet adhesion and aggregation, and forming thrombosis^12, 26^, which may increase the risk of adverse clinical outcomes after ischemic stroke. Our findings support the above-mentioned relationship between HS-CRP and platelets and suggest that there is an effect of the interaction between HS-CRP and platelet count on adverse clinical outcomes after ischemic stroke.

Antiplatelet therapy is an important component of the secondary prevention of ischemic stroke^36^; therefore, we adjusted for antiplatelet therapy in a multivariable model as an important confounder. Thus, the effect of antiplatelet therapy on the credibility of the present results is minimal. At present, there are no RCTs on anti-inflammatory drugs for ischemic stroke prognosis, but Ridker et al.^42^ found that the use of statins reduced the incidence of stroke and all-cause mortality in a healthy population with elevated HS-CRP levels, but without hyperlipidemia. This provides a perspective on the use of statins to reduce inflammation in patients with stroke.

Laboratory testing for the platelet count and HS-CRP is universal and inexpensive, and the combined detection of the two indices can better predict the prognosis of ischemic stroke, and our findings suggest that antiplatelet and anti-inflammatory therapies should be taken simultaneously when the two indices rise. Therefore, our findings have important clinical implications not only for improving the risk stratification of ischemic stroke, but also for suggesting a combination of both antiplatelet and anti-inflammatory therapies in ischemic stroke patients, especially in patients with elevated platelet and HS-CRP levels.

Our study has several limitations. First, patients with BP ≥ 220/120 mmHg or severe cardiovascular disease or those undergoing intravenous thrombolytic therapy were excluded due to the CATIS design; thus, this study might have some selection bias. However, the proportion of patients with BP ≥ 220/120 mmHg or those treated with intravenous thrombolytic therapy is low in China^27, 43^, and the baseline characteristics of participants in this study were similar to those from the China National Stroke Registry^44^, suggesting that the selection bias may be minimal.

Second, we only collected data on serum HS-CRP levels and platelet counts at admission, not their follow-up data; thus, the association between the changes in these biomarkers and stroke prognosis cannot be estimated. Third, this was an observational study; thus, the possibility of residual confounding could not be fully eliminated, although we did adjust for almost all of the important confounders.

## Conclusion

We found that the prognostic value of the platelet count was modified by HS-CRP in patients with ischemic stroke. Elevated platelet counts were associated with adverse clinical outcomes only in ischemic stroke patients with high HS-CRP levels, suggesting that risk stratification of ischemic stroke patients should be conducted and strategies for anti-inflammatory and antiplatelet therapy be developed according to the results of both platelet and HS-CRP testing. Further prospective studies from other populations and randomized clinical trials are required to verify our findings.

## Data Availability

All data included in this study are available upon request by contact with the corresponding author.

## Abbreviations

CATIS: China Antihypertensive Trial in Acute Ischemic Stroke
HS-CRP: high-sensitivity C-reactive protein
BP: blood pressure
NIHSS: National Institutes of Health Stroke Scale
mRS score: modified Rankin Scale
(ALLHAT): Antihypertensive and Lipid-Lowering Treatment to Prevent Heart Attack Trial
OR: odds ratio
HR: hazard ratio
CI: confidence intervals
BMI: body mass index
SBP: systolic blood pressure
DBP: diastolic blood pressure
TG: triglycerides
TC: total cholesterol
LDL-C: low density lipoprotein cholesterol
HDL-C: high density lipoprotein cholesterol
TIA: transient ischemic attack

## Acknowledgments

We thank the study participants, their relatives, and the clinical staff at all participating hospitals for their support and contributions to this project.

## Authors’ contributions

Fanghua Liu and Pinni Yang wrote the main manuscript text. Fanghua Liu and Yonghong Zhang were contributed to the idea of the study. Yinan Wang, Mengyao Shi, Ruirui Wang, Qingyun Xu, Yanbo Peng, Jing Chen, Jintao Zhang, Aili Wang, Tan Xu, Yonghong Zhang and Jiang He were contributed to the collection of data and the statistical analysis. All authors read and approved the final manuscript.

## Availability of data and materials

Not applicable.

## Ethics approval and consent to participate

Not applicable.

## Consent for publication

Not applicable.

